# Olfactory and Gustatory Dysfunction as An Early Identifier of COVID-19 in Adults and Children: An International Multicenter Study

**DOI:** 10.1101/2020.05.13.20100198

**Authors:** Chenghao Qiu, Chong Cui, Charlotte Hautefort, Antje Haehner, Jun Zhao, Qi Yao, Hui Zeng, Eric J. Nisenbaum, Li Liu, Yu Zhao, Di Zhang, Corinna G. Levine, Ivette Cejas, Qi Dai, Mei Zeng, Philippe Herman, Clement Jourdaine, Katja de With, Julia Draf, Bing Chen, Dushyantha T. Jayaweera, James C. Denneny, Roy Casiano, Hongmeng Yu, Adrien A. Eshraghi, Thomas Hummel, Xuezhong Liu, Yilai Shu, Hongzhou Lu

**Affiliations:** Center of Stomatology, Shanghai Public Health Clinical Center, Shanghai 201508, China; ENT Institute and Otorhinolaryngology Department of the Affiliated Eye and ENT Hospital, State Key Laboratory of Medical Neurobiology, Institutes of Biomedical Sciences, Fudan University, Shanghai, 200031, China; NHC Key Laboratory of Hearing Medicine, Fudan University, Shanghai, 200031, China; Department of Head and Neck Surgery,Hopital Lariboisiere, University of Paris, Paris, France; Smell and Taste Clinic, Department of Otorhinolaryngology, TU Dresden, Dresden, Germany; Center of Pediatrics, Shanghai Public Health Clinical Center, Shanghai 201508, China; Department of Otorhinolaryngology, Chinese and Western Medicine Hospital of Tongji Medical College, Huazhong University of Science and Technology, Wuhan, 430022, China; Department of Cardiovascularology, The Third People’s Hospital of Shenzhen 29 Bulan Road, Longgang District, Shenzhen, 518112, China; Department of Otolaryngology, University of Miami Miller School of Medicine, Miami, FL 33136, USA; Department of Infectious Diseases, Shanghai Public Health Clinical Center, 2901 Caolang Road, Shanghai 201508, China; Department of Otolaryngology, The Third People’s Hospital of Shenzhen, 29 Bulan Road, Longgang District, Shenzhen, 518112, China; Department of Infectious Diseases, Children’s Hospital of Fudan University 399 Wanyuan Road, Shanghai, China, 201102; Division of Infectious Diseases, University Hospital Carl Gustav Carus at the TU Dresden, Dresden, Germany; Department of Medicine, Division of Infectious Diseases, University of Miami Miller School of Medicine, Miami, FL 33136, USA; American Academy of Otolaryngology—Head and Neck Surgery, Alexandria, Virginia, USA

**Author notes:** contributed equally (Chenghao Qiu, Chong Cui, Charlotte Hautefort, Antje Haehner, Jun Zhao, Qi Yao, Hui Zeng, Eric J. Nisenbaum). **Corresponding Author:** Dr. Xuezhong Liu, Department of Otolaryngology (D-48), University of Miami, 1666 NW 12th Avenue, Miami, Florida 33136, USA, Dr. Yilai Shu, ENT Institute and Otorhinolaryngology Department of the Affiliated Eye and ENT Hospital, State Key Laboratory of Medical Neurobiology, Fudan University, Shanghai, China., Dr. Thomas Hummel, Smell and Taste Clinic, Department of Otorhinolaryngology, TU Dresden, Dresden, Germany, Dr. Hongzhou Lu Department of Infectious Diseases, Shanghai Public Health Clinical Center, 2901 Caolang Road, Shanghai 201508, China.

**Keywords:** COVID-19, SARS-CoV-2, anosmia, dysgeusia, COVID-19 screening, olfactory dysfunction, gustatory dysfunction

## Abstract

**Objective:** Evaluate the prevalence and characteristics of olfactory or gustatory dysfunction in COVID-19 patients

**Study Design:** Multicenter Case Series

**Setting:** 5 tertiary care hospitals (3 in China, 1 in France, 1 in Germany)

**Subjects and Methods:** 394 PCR confirmed COVID-19 positive patients were screened, and those with olfactory or gustatory dysfunction were included. Data including demographics, COVID-19 severity, patient outcome, and the incidence and degree of olfactory and/or gustatory dysfunction were collected and analyzed. The Questionnaire of Olfactory Disorders (QOD) and Visual Analogue Scale (VAS) were used to quantify olfactory and gustatory dysfunction respectively. All subjects at one hospital (Shanghai) without subjective olfactory complaints underwent objective testing.

**Results:** Of 394 screened subjects, 161 (41%) reported olfactory and/or gustatory dysfunction and were included. Incidence of olfactory and/or gustatory disorders in Chinese (n=239), German (n=39) and French (n=116) cohorts were 32%, 69%, and 49% respectively. The median age of included subjects was 39 years old, 92/161 (57%) were male, and 10/161 (6%) were children. Of included subjects, 10% had only olfactory or gustatory symptoms, and 19% had olfactory and/or gustatory complaints prior to any other COVID-19 symptom. Of subjects with objective olfactory testing, 10/90 demonstrated abnormal chemosensory function despite reporting normal subjective olfaction. 43% (44/102) of subjects with follow-up showed symptomatic improvement in olfaction or gustation.

**Conclusions:** Olfactory and/or gustatory disorders may represent early or isolated symptoms of SARS-CoV-2 infection. They may serve as a useful additional screening criterion, particularly for the identification of patients in the early stages of infection.

## Introduction

The global spread of SARS-CoV-2 is fast-moving and ongoing, with more than three million cases reported across 214 countries and regions.^1^ The main clinical symptoms of COVID-19 patients are fever, dry cough, and fatigue. Patients may also present with less typical symptoms including nasal congestion, rhinorrhea, sore throat, myalgia, and diarrhea.^2^ Recently reports from a number of countries indicate that olfactory or gustatory dysfunction may also be one of the initial presenting symptoms in a large number of COVID-19 patients.^3^ A study of COVID-19 inpatients in Wuhan demonstrated that 5.1% (11/214) patients experienced hyposmia, and 5.6% (12/214) patients exhibited hypogeusia.^4^ An Italian study found that 33.9% of COVID-19 confirmed inpatients presented with at least one olfactory or gustatory symptom.^5^ A study of patients with influenza-like symptoms showed that COVID-19 positive patients have a high prevalence of olfactory or gustatory disorders compared with COVID-19 negative patients.^6^ Olfactory and gustatory disorders were also found to be prevalent symptoms in COVID-19 patients in a multicenter European study.^7^ The American Academy of Otolaryngology-Head and Neck Surgery (AAO-HNS) developed an anosmia reporting tool that was opened to contributors on March 26,2020 and the initial findings were reported on 237 patients worldwide. There are now over 800 patients submitted through this tool. Anosmia was the sentinel symptom in over 25% of the patients and occurred in over two-thirds of the patients.^8^ Some patients with mild symptoms may only have changes in olfaction or gustation and so may be overlooked in screening and testing efforts, leading to further disease spread.

However, the documented prevalence of olfactory or gustatory disorders differs greatly between studies, and little is known about how this may vary based on factors such as age, disease severity, and between multiple different countries and/or ethnic populations. To remedy this, we studied the association between olfactory and gustatory complaints and early SARS-CoV-2 infections at 5 hospitals in China, Germany and France. This study aims to systematically characterize and compare olfactory and gustatory symptoms among COVID-19 adult patients and children in five epidemic areas in Shanghai, Wuhan and Shenzhen [China], Paris [France] and Dresden [Germany], emphasizing the importance of these symptoms as an early marker of SARS-CoV-2 infection.

## Method

Through retrospective chart review, 394 COVID-19 patients presenting from March 15th, 2020 to April 5th, 2020 (141 cases in Shanghai, China; 62 cases in Wuhan, China; 36 cases in Shenzhen, China; 39 cases in Dresden, Germany; and 116 cases in Paris, France) initially diagnosed by RT-PCR from oro- or nasopharyngeal swab specimens were screened. All 394 patients were asked about loss of olfaction and/or gustation and 161 patients presenting with olfactory and/or gustatory dysfunction were identified and enrolled in this study, with final follow-up obtained on April 5th, 2020. Hospitals included in the study were as follows: Shanghai Public Health Clinical Center of Fudan University, China (61 patients including 9 adolescents), Chinese and Western Medicine Hospital of Tongji Medical College, Huazhong University of Science and Technology, China (11 patients), Shenzhen Third People’s Hospital, China (5 patients), Lariboisière University Hospital, France (57 patients) and University Hospital at the TU Dresden, Germany (27 patients including one adolescent). The three Chinese hospitals are designated by the government for the treatment of COVID-19 patients. A visual analogue scale (VAS) of olfactory intensity and questionnaire of olfactory disorders (QOD) were used to assess the presence and degree of hyposmia. The QOD-P(parosmic statements) evaluates the characteristic of the olfactory disorder, with a high score indicating the presence of an olfactory disorder; QOD-QoL(quality of life) evaluates quality of life, with a high score indicating a serious olfactory impairment; QOD-DS(socially desired statements) evaluates reporter reliability, with a low score indicating a tendency to give answers in line with perceived social expectations.^9^ All COVID-19 patients in Shanghai, China without subjective olfactory complaints underwent objective olfactory testing with the following stimuli to screen for the presence of subclinical hyposmia: eugenol (the smell of cloves – olfactory stimulus), 75% ethanol (trigeminal stimulus), and vinegar (mixed olfactory-trigeminal stimulus). For the test, three disposable cotton swabs dipped in each of these stimuli were placed 1-2 cm below the front nostril of the subjects, who were instructed to smell 3 times. A VAS of gustatory intensity was used to assess the presence and degree of hypogeusia.

Disease severity was classified as follows. Asymptomatic patients indicates that there were no clinically identifiable or self-perceived signs or symptoms of infection at the time of positive SARS-CoV-2 PCR testing. Patients classified as mild presented with only low fever, mild cough, slight fatigue, and no evidence of pneumonia on CT imaging. Patients classified as moderate had fever and respiratory symptoms, and imaging showed evidence of pneumonia. Patients classified as severe had dyspnea, respiratory frequency ≥30/min, blood oxygen saturation ≤93%, partial pressure of arterial oxygen to fraction of inspired oxygen ratio ≤300mmHg or pulmonary CT imaging showing at least a 50% increase in infiltrate volume over 24-48 hours. Patients classified as critical had respiratory failure, septic shock, and/or multiple organ dysfunction or failure.^2^

Clinical, treatment, and outcome data were obtained from data collection forms in the electronic medical records. The data were analyzed and reviewed by a team of trained physicians. The recorded information included demographic data, comorbidities, severity classification, patient outcomes, the degree of olfactory or gustatory loss and the incidence of the symptom in COVID-19 patients. A descriptive analysis was performed. Categorical variables were described as frequency rates and percentages, and continuous variables were described using mean, median, standard deviation (SD) and interquartile range (IQR) values. A pairwise comparison of data from the three countries was made to assess for demographic heterogeneity and examine differences in the prevalence and characteristics of olfactory and gustatory symptoms. Independent group t tests and chi-square tests were used for continuous variables and categorical variables, respectively. Fisher exact test was used when the data were limited. All statistical analyses were performed using SPSS (Statistical Package for the Social Sciences) version 26.0 software (SPSS Inc). Statistical significance was set at a 2-sided α of less than 0.05. Percentages were rounded to the nearest whole number. This study was approved by the Institutional Ethics Committees of Shenzhen Third People’s Hospital, Wuhan Chinese and Western Medicine Hospital, and Shanghai Public Health Clinical Center; the Ethics Committee of the University Clinic, TU Dresden; and the Ethics Committee of Lariboisière University Hospital.

## Results

Of the 394 COVID-19 patients screened for the study, 161 (41%) exhibited sudden olfactory and/or gustatory dysfunction and were included. The incidence of olfactory and/or gustatory complaints in COVID-19 patients in China, Germany and France is 32%, 69% and 49%, respectively (China VS Germany, P<0.001; China VS France, P=0.002; Germany VS France, P=0.029). Sixty-one of 394 (15 %) of patients reported isolated olfactory dysfunction and 7 of 394 (2%) reported isolated gustatory dysfunction. Ninety-three of 394 (24%) patients had both olfactory and gustatory dysfunction. Classification of COVID-19 severity was available for 278 observed patient in China and Germany. The incidence of olfactory or taste dysfunction in mild, moderate and severely ill patients was 53/98 (54%), 40/107 (37%) and 11/63 (17%),respectively. The severity classification among all COVID-19 patients in France was unavailable.

Among the 161 included patients who had sudden olfactory or gustatory loss, median age was 38.8±17.6 years (IQR, 23-53 years) and 92/161 (57%) were male. Distribution of age in these enrolled patients was significantly different between China and the two European countries (China VS Germany, P=0.001; China VS France, P < 0.001; Germany VS France, P=0.165). There were no statistically significant differences in gender among the three countries (China VS Germany, P=0.354; China VS France, P=0.888; Germany VS France, P=0.321). Overall, seventy-eight of 161 (48%), 40/161 (25%), 33/161 (20%) and 10/161 (6%) were in mild, moderate, severe and critical condition, respectively. However, distribution of disease severity in patients with olfactory or gustatory dysfunction varied significantly between countries (China VS Germany, P<0.001; China VS France, P<0.001; Germany VS France P<0.001). Of the 161 patients, 156 were able to provide information on the timing and order of symptom onset. Overall, 15/156 (10%) patients had olfactory or gustatory dysfunction as their only symptom of COVID-19 infection. The percentage of patients with olfactory or gustatory disorders as their only symptom in China, Germany and France were 12%, 4% and 9%, respectively. Twenty-nine of 156 (19%) patients experienced olfactory or gustatory dysfunction as the first symptom of their SARS-CoV-2 infection. The percentage of patients presenting with olfactory or gustatory disorders as their first symptom of infection in China, Germany and France were 23%, 15% and 15%, respectively. Olfactory or gustatory symptoms occurred simultaneously with classic COVID-19 symptoms such as fever, cough, sore throat and diarrhea in 30 of 156 (19%) patients (Table 1). A single patient developed hearing loss during the course of their COVID-19 infection.

**Table 1.** Characteristics of COVID-19 Patients Presenting with Olfactory/Gustatory Dysfunction.

| Characteristics | Countries |  |  | Total |
| --- | --- | --- | --- | --- |
|  | China | Germany | France |  |
| Olfactory/gustatory dysfunction<br>No./Total screened No.(%) | 77/239 (32%) | 27/39 (69%) | 57/116 (49%) | 161/394 (41%) |
| Symptoms,<br>No/Total screened No.(%) |  |  |  |  |
| Only olfactory dysfunction | 47/239 (20%) | 7/39 (18%) | 7/116 (6%) | 61/394 (15%) |
| Only gustatory dysfunction | 6/239 (3%) | 1/39 (3%) | 0/116 (0) | 7/394 (2%) |
| Olfactory and gustatory dysfunctions | 24/239 (10%) | 19/39 (49%) | 50/116 (43%) | 93/394 (24%) |
| Age of enrolled subjects, median ± SD (IQR) | 30.5 ± 16.2 (20, 35) | 43.1 ± 15.2 (32, 55.5) | 48.1 ± 15.3 (33, 60) | 38.8 ± 17.6 (23, 53) |
| Sex of enrolled subjects, No/Total enrolled No.(%) |  |  |  |  |
| Male | 45/77 (58%) | 13/27 (48%) | 34/57 (60%) | 92/161 (57%) |
| Female | 32/77 (42%) | 14/27 (52%) | 23/57 (40%) | 69/161 (43%) |
| COVID-19 severity in enrolled subjects No/Total enrolled No.(%) |  |  |  |  |
| Asymptomatic | 0/77 (0) | 0/27 (0) | 0/57 (0) | 0/161 (0) |
| Mild | 26/77 (34%) | 27/27 (100%) | 25/57 (44%) | 78/161 (48%) |
| Moderate | 40/77 (52%) | 0/27 (0) | 0/57 (0) | 40/161 (25%) |
| Severe | 11/77 (14%) | 0/27 (0) | 22/57 (39%) | 33/161 (20%) |
| Critical | 0/77 (0) | 0/27 (0) | 10/57 (18%) | 10/161 (6%) |
| Onset of olfactory/gustatory dysfunction, No/Total enrolled No.(%) |  |  |  |  |
| As the only symptom of COVID-19 | 9/75 (12%)<sup>a</sup> | 1/27 (4%) | 5/54 (9%)<sup>b</sup> | 15/156 (10%)<sup>a,b</sup> |
| Before general symptoms of COVID-19 | 17/75 (23%)<sup>a</sup> | 4/27 (15%) | 8/54 (15%)<sup>b</sup> | 29/156 (19%)<sup>a,b</sup> |
| At the same time as general symptoms of COVID-19 | 5/75 (7%)<sup>a</sup> | 4/27 (15%) | 21/54 (39%)<sup>b</sup> | 30/156 (19%)<sup>a,b</sup> |
| After general symptoms of COVID-19 | 44/75 (59%)<sup>a</sup> | 18/27 (67%) | 20/54 (37%)<sup>b</sup> | 82/156 (53%)<sup>a,b</sup> |
| Outcome of olfactory/gustatory Symptoms, No/ Total enrolled No.(%) |  |  |  |  |
| Improved | 34/77 (44%) | Unknown/27<sup>c</sup> | 10/25 (40%)<sup>d</sup> | 44/102 (43%)<sup>f</sup> |
| Non-improved | 43/77 (56%) | Unknown/27<sup>c</sup> | 15/25 (60%)<sup>d,e</sup> | 58/102 (57%)<sup>f</sup> |

Abbreviations: SD, standard deviation; IQR, interquartile range. <sup>a</sup>two patients in China could not remember the date of onset of olfactory or gustatory dysfunction.
<sup>b</sup>three patients in France could not remember the date of onset of olfactory or gustatory dysfunction. <sup>c</sup>the patients' improvement in Germany is unavailable. <sup>d</sup>only 25 patients were followed in France. <sup>e</sup>three critical patients died of COVID-19. <sup>f</sup>statistics for China(77 cases) and France(25 cases).

In a subset of patients we were able to evaluate olfactory or gustatory function using the olfactory VAS (n=113), the gustatory VAS (n=46) and the QOD (n=60). The mean VAS scores of patients with olfactory or gustatory dysfunction were 3.60±3.62 (IQR 0-7) and 4.46±2.90 (IQR 2.25-6), respectively. The mean scores of QOD-QoL (quality of life) were 37%±23% (IQR 18%-49%), QOD-DS (socially desired statements) were 41%±15% (IQR 32%-50%), and QOD-P (parosmic statements) were 40%±30% (IQR 17%–60%) Table 2). Of the 90 patients without subjective olfactory dysfunction in the Shanghai, China cohort who were tested with the three chemosensory stimuli, 10 (9 adults and 1 child) showed abnormal chemosensory function. Eight of the ten patients could not smell the three stimuli at all, while the other two could smell an odor but could not distinguish the type of odor. These eight patients who could not perceive vinegar and 75% ethanol may also have a trigeminal deficit.

**Table 2.** The Scores of VAS and QOD in COVID-19 Patients Presenting with Olfactory/Gustatory Dysfunction.

| Score | Symptoms |  |
| --- | --- | --- |
|  | Olfactory Dysfunction | Gustatory Dysfunction |
| VAS, median±SD (IQR) | 3.60±3.62 (0, 7) | 4.46±2.90 (2.25, 6) |
| QOD, median (IQR) |  |  |
| QoL | 37%±23%(18%, 49%) | - |
| DS | 41%±15%(32%, 50%) | - |
| P | 40%±30%(17%, 60%) | - |

Abbreviations: VAS, visual analogue scale; QOD, the questionnaire of olfactory disorders; SD, standard deviation; IQR, interquartile range; QoL, quality of life; DS, socially desired statements; P, parosmic statements.

Of the 102 patients for whom follow-up data was available (time range for follow-up was 3 weeks), 44 of 102 (43%) patients had improvement of their olfactory or gustatory dysfunction, with the other 58 (57%) showing no improvement (including 3 deaths among the critical cases). There was no significant difference in the outcome of olfactory and/or gustatory disorders between Chinese and French COVID-19 patients (P=0.715). Follow-up data was not available for the German cohort, and was available for only 25 patients in the French cohort (Table 1).

Of 27 COVID-19 positive children who were screened (range of age 6-17 years), 10 (37%) children with olfactory or gustatory dysfunction were included. The age of these 10 patients ranged from 15 to 17 years old, and six of 10 (60%) were male. Seven children (3 mild cases, 4 moderate cases) had both hyposmia and hypogeusia. Only eight of the 10 patients were able to report the sequencing of their symptoms. Of these eight, two patients (25%) had hyposmia and/or hypogeusia as their only symptom, and both were found to have infected their family member prior to diagnosis. An additional hospitalized child who did not report any subjective olfactory symptoms was also found to have subclinical olfactory dysfunction when tested with the olfactory reagents. Three of 9 (33%) children showed total symptomatic improvement during the observation period. Follow-up was not obtained for one child in the German cohort (Table 3).

**Table 3.** Characteristics of COVID-19 Positive Children Presenting with Olfactory/Gustatory Dysfunction.

| Characteristics | Children |
| --- | --- |
| Total enrolled No./Total observed No.(%) | 10/27 (37%) |
| Age, median±SD(IQR),y | 16.6±0.7 (16.3, 17) |
| Sex, No/Total enrolled No.(%) |  |
| Male | 6/10 (60%) |
| Female | 4/10 (40%) |
| COVID-19 severity classification, No/Total enrolled No.(%) |  |
| Asymptomatic | 0/10 (0) |
| Mild | 6/10 (60%) |
| Moderate | 4/10 (40%) |
| Severe | 0/10 (0) |
| Critical | 0/10 (0) |
| Symptoms, No/ Total observed No.(%) |  |
| Only olfactory dysfunction | 3/27 (11%) |
| Only gustatory dysfunction | 0/10 (0) |
| Olfactory and gustatory dysfunctions | 7/27 (26%) |
| Onset of olfactory/gustatory dysfunction, No/ Total enrolled No.(%) |  |
| As the only symptom of COVID-19 | 2/8 (25%)<sup>a</sup> |
| Before general symptoms of COVID-19 | 0/8 (0)<sup>a</sup> |
| At the same time as general symptoms of COVID-19 | 1/8 (13%)<sup>a</sup> |
| After general symptoms of COVID-19 | 5/8 (63%)<sup>a</sup> |
| Outcome of Olfactory/Gustatory Symptom, No/Total enrolled No.(%) |  |
| Improved | 3/9 (33%)<sup>b</sup> |
| Non-improved | 6/9 (67%)<sup>b</sup> |

Abbreviations: SD, standard deviation; IQR, interquartile range <sup>a</sup> two children could not remember the date of onset of olfactory or gustatory dysfunction. <sup>b</sup>follow-up was not obtained for one child in the German cohort.

## Discussion

This study systematically studied the incidence and characteristics of sensory dysfunction among COVID-19 patients. It is the first to examine olfactory and/or gustatory dysfunction in infected children. In our cohort of 394 COVID-19 patients, 41% presented with olfactory and/or gustatory disorders, of whom approximately half were mildly symptomatic. The incidence of olfactory or gustatory disorders in COVID-19 patients in Germany (69%) and France (49%) were significantly higher than in China (32%, P<0.001 and P=0.002 respectively). A recent study also found that different variants of SARS-CoV-2 are predominant outside of East Asia (“Type A” and “Type C”) compared to within East Asia (“Type C”).^10^ It is possible that phenotypic characteristics – including incidence of olfactory and gustatory disorders – may differ between these variants. There is also the possibility that the incidence of olfactory or gustatory disorders may vary based on ethnicity. However, given that there was also a smaller but significant different between the German and French groups (P=0.029), it is possible that the differences reflect other sources of heterogeneity between the groups such as subject age, as the median age of German and French groups showed significant difference with Chinese group (P=0.001 and P<0.001 respectively). Interestingly, the distribution of COVID-19 disease severity among included subjects varied significantly when comparing the three country cohorts to each other, which may indicate that presence of olfactory or gustatory symptoms are not related to overall disease severity. Olfactory or gustatory symptoms were the first and only symptoms in 10% of patients, and 19% of patients experienced olfactory or gustatory dysfunction prior to any other COVID-19 symptoms. In addition, 25% of children had only olfactory or gustatory symptoms at COVID-19 presentation. This indicates that olfactory or gustatory disorders are early identifiers of COVID-19 in a subset of patients, and thus has important implications for patient screening and disease control.

Despite still being infectious, a significant portion of COVID-19 patients may be minimally or atypically symptomatic early in the course of the disease, with one study finding that less than 50% of early patients are febrile.^11^ These patients may be missed by screening initiatives focused on “classic” symptoms such as fever and cough, delaying diagnosis, undermining triage and quarantine efforts, and leading to higher rates of disease transmission. At least two mild patients in our study who presented with isolated olfactory dysfunction are documented to have infected others prior to admission despite the robust screening and triage measures implemented at the time, highlighting the potential danger. Adding questions about olfactory or gustatory changes to routine screening measures would help identify these patients earlier, a boon to infection control initiatives. Due to the inherent difficulties of COVID-19 patient coordination, it is hard to perform T & T olfactometer test, though disposable olfactory tests like the University of Pennsylvania Smell Identification Test (UPSIT) could be applied. Notably, there were 10 subjects (9 adults and 1 child) who reported that they had no olfactory dysfunction, but demonstrated signs of subclinical hyposmia when exposed to three chemosensory stimuli. This may suggest that screening based on self-reported symptoms alone is not sufficient, considering the relatively high degree of erroneous self-ratings with respect to the chemical senses.^12^ However, it is not certain if any of these patients had unrecognized pre-existing smell and/or taste disorders unrelated to Covid-19.

Recent studies have shed light on the mechanisms that may underlie the loss of olfaction or gustation. SARS-CoV-2 has been found to replicate particularly well in the nose, with high viral loads detected there soon after symptom onset.^13^ ACE2 - the main host cell receptor of SARS-CoV-2 – and TMPRSS2 – a cell surface protease involved in SARS-CoV-2 cell entry - are highly expressed in a variety of olfactory epithelial cell types, raising the possibility that viral invasion of these cells may lead to anosmia.^14,15^ It has also been suggested that transcribiform viral spread and infection of more proximal elements of the olfactory pathway via CNS ACE2 receptors may also contribute to olfactory dysfunction.^16,17^ Alternatively, it is possible that at least some of the olfactory dysfunction is conductive – secondary to nasal congestion, swelling, or inflammation preventing olfactory molecules from reaching the olfactory cleft – rather than sensorineural.^18^ This may be particularly true in the subset of patients who showed improvement of their olfactory dysfunction on follow-up, though further research is needed. ACE2 is also highly expressed on the oral mucosa and tongue, representing a potential mechanism for gustatory dysfunction.^19^

In addition to the much more common olfactory and gustatory sensory deficits, we identified new onset hearing loss in one severely ill patient. While more research is needed, this suggests that we should also be alert for other sensory deficits in addition to hyposmia and dysgeusia, particularly given SARS-CoV-2’s ability to invade the CNS.

## Limitations

As with any retrospective study, the conclusions that can be drawn are limited by the heterogeneous nature of the data available. Likewise, due to the impact of the COVID-19 pandemic and the difficulties inherent to a retrospective multi-institutional and international study, there are differences in data collection methods and patient populations between institutions that make drawing direct comparisons for some measures more difficult. Olfactory and gustatory function have not been measured psychophysically in COVID-19 positive patients, so it is possible that patients may have confused gustatory and retronasal olfactory function, which would result in biased ratings of olfactory or gustatory function. In addition, the subjective perception of olfactory function is confounded by nasal breathing. Finally, symptom timeline often relies on patient report of their symptoms and is subject to recall bias. To address these issues, future prospective studies are necessary.

## Conclusion

Olfactory and/or gustatory dysfunction may represent early or even the only symptom of SARS-CoV-2 infection in both adults and children. As such, they may serve as important screening criteria for identifying otherwise oligosymptomatic or atypically symptomatic patients missed by other screening measures. We recommend that questions related to olfactory and gustatory changes be added to routine screening measures to help identify these patients prior to further disease transmission.

## Data Availability

The data for the present study are available from the corresponding author upon reasonable request.

## Author Contributions

Yilai Shu, Xuezhong Liu, Thomas Hummel and Hongzhou Lu contributions to conception and design, interpretation of data, drafting the article, final approval of the version to be published; Chong Cui, Yilai Shu, Chenghao Qiu, Charlotte Hautefort,Antje Haehner, Jun Zhao, Qi Yao, Hui Zen, Li Liu, Mei Zeng contributions to acquisition of data, analysis and interpretation of data, and design; Eric Nisenbaum, Yu Zhao, Di Zhang contributions to interpretation of data, writing, revising it critically for important intellectual content, final approval of the version to be published; Philippe Herman, Clement Jourdain contributions to acquisition of data; Corinna Levine, Ivette Cejas, Qi Dai, Katja de With, Julia Draf, Bing Chen, Dushyantha T. Jayaweera, James C. Denneny III, Roy Casiano, Hongmeng Yu, Adrien A. Eshraghi interpretation of data, writing, revising it critically. All of them agreement to be accountable for all aspects of the work in ensuring that questions related to the accuracy or integrity of any part of the work are appropriately investigated and resolved.

